# Digital quantification of chronic gastric wall inflammation in asymptomatic bariatric patients after laparoscopic sleeve gastrectomy

**DOI:** 10.1101/2020.02.11.20022160

**Authors:** Camelia D Vrabie, Bogdan Geavlete, Marius Gangal

**Author notes:** Corresponding author: Marius Gangal MD, PhD, MBA, Medical Data Analytics Canada (MEDACS.CA), Rigaud, J0P1P0, PQ Canada.

## Abstract

**Aim:** to evaluate validity of digital quantification when compared with human fast-scoring in routine classification of chronic gastric wall inflammation.

**Method:** 87 bariatric gastric samples coming from asymptomatic severe obese patients were examined and classified as normal, with unspecified chronic gastritis and lymphocytic gastritis using a fast-scoring, visual analogue scale method. Results were compared with digital diagnostic data (manual segmentation, supervised learning analysis based on intraepithelial lymphocytes count criteria). Discordant results were re-evaluated by the human pathologist by direct count (ground truth). Helicobacter Pylori diagnostic was performed in all cases (Giemsa).

**Results:** Digital analysis classified chronic inflammation as lymphocytic gastritis in 45 cases (mean 53 lymphocytes / 100 epithelial gastric cells ±18). 30 cases were labeled as unspecific chronic gastritis (mean 25/100±2.8) (p<0.0001). Human fast-scoring classified 43 cases as lymphocytic gastritis and 20 as unspecific gastritis. Helicobacter Pylori was detected in 49% of lymphocytic gastritis cases and in 7% of chronic gastritis. 47 diagnostics were concordant (54%). In 36%, digital score was better than human fast-scoring. In 7%, digital results were false negative (all cases generated by technical artifacts). Overall, digital quantification had 89% accuracy and 96% precision when compared with ground truth.

**Conclusion:** In our study, digital image analysis produced a fast and reproducible classification of chronic gastric inflammation with good precision and accuracy. Technical errors generated 6 cases of false negative results. Several other limitations of the study (use of only bariatric gastric fundus tissue, low number of cases, manual supervised learning segmentation) ask for an increased number of cases evaluation before validation and clinical use.

## Introduction

Chronic Gastric inflammation (Chronic gastritis - CG) is a common histologic finding in many medical conditions (1). It is often associated with the presence of Helicobacter Pylori (HPyl), an opportunistic gram-negative bacterium that infects more than 50%CV and BG conceived the idea, CV5 of world population (2). A correlation between HPyl infestation and the development of CG, peptic ulcer, gastric adenocarcinoma and gastric lymphoma was often reported (3,4). Bacterial induced CG has histological features (mononuclear cellular infiltrate that can be associated with active inflammation) (5) that allow direct visual grading and classification of inflammatory cells. As the process is laborious and time-consuming, it is not sustainable in the daily routine activity. A four points visual analogue scale was adopted for fast grading (6) and is frequently used in routine histopathology exams (RHP). RHP is a microscopic exam performed under standard conditions on any surgical tissue, even if there is minimal or no suspicion of a clinically significant abnormality (7). It is supposed to offer a fast and accurate diagnostic that will improve the management of patients after any surgery but is not always precise, fast or cost-effective, at least after some interventions (8). More challenging, when RHP is evaluating randomly sampled, normal specimens, the process is labor intense, time and budget consuming without producing significant results (9,10). RHP can be a medico-legal requirement. It is often mandated by health laws or regulations and more than 50% of any average surgical pathology lab time and budget is dedicated to it (11). There are authors that asked for a more strategic use of RHP (12) but the decision is not easy as RHP may detect serious incidental findings that can influence the treatment of any surgical patient (13).

RHP after bariatric surgery reflects well the dilemma of routine histology performed on normal tissue excised for therapeutic reasons. Obesity is an important global public health problem of epidemic proportions with a multifactorial pathogenesis that may generate serious complications. Between many existing treatment options, bariatric surgical procedures gained popularity. Laparoscopic Gastric Sleeve (LGS) is a recently added surgical technique able to control both the disease and associated comorbidities in a safe and efficacious way (14) by removing a part of the “normal” gastric wall (15). As obesity does not alter the structure of the gastric wall, in theory, surgical pathology diagnostics after bariatric interventions should report only normal or incidental findings (16). In reality, the number and seriousness of pathology diagnostics after bariatric surgery is substantial (17) mainly because of Helicobacter Pylori (HPyl) gastric infection. As after any gastric surgery, HPyl may also generate early post bariatric complications (18) fact that makes diagnostic and treatment of HPyl infestation, a priority (19).

In bariatric patients that do not have any gastric symptomatology, chronic lymphocytic inflammation diagnostic is performed mainly during the RHP exam. The use of a four points Visual Analog Scale (VAS) (classifying the intraepithelial infiltrate as normal, mild, moderate or severe) (20) makes the process fast, but subjective.

A possible solution to improve routine CG classification is provided by digital pathology. Digital pathology research started in the mid of twentieth century (21) and gained credibility once computer power increased and relevant software was developed. Within the field of digital pathology, digital image analysis (DIA) proved to be a useful diagnostic tool, able to reduce diagnostic subjectivity, allowing remote consultation possibilities, better training and lab resources use (22,23). Even not entirely validated outside research, DIA showed excellent results in diagnosing several forms of breast and colon cancers (24). In benign histology, tissue characteristics (including stromal differentiation and possible post-harvest tissue necrosis), associated disorders, sampling and staining errors may add challenges to DIA diagnostic performance (25). Any digital analysis workflow will involve several steps: image data acquisition and ground truth generation, image analysis including object detection, segmentation and recognition, and medical statistics (26). All of these steps can be performed manually with the help of open-source software (adapted for specific laboratory needs) or can be automated, involving the use of whole slide scanners and proprietary software. Manual acquisition and analysis of microscopic slides is more economically affordable but can be as subjective as human reading. Machine scanners can produce precise histologic diagnostics (27) but are still expensive as a solid informatics network is additionally required (28).

Our research objective was to evaluate how manual DIA may improve routine (HE) chronic gastric classification compared to human fast-scoring in tissue sampled from asymptomatic bariatric patients that had an LGS intervention.

## Materials and Methods

One surgical pathologist examined all gastric samples from consecutive bariatric patients that had LGS surgery (29) over one year (87 cases). Six fragments of gastric mucosa were sampled and prepared for routine histology. One 4-microns section was taken from each block and stained with HE then examined with a standard optical microscope (objective ×10, camera 12MP). Derived from Sydney criteria (20), gastric tissue was scored as normal, with mild, moderate or severe inflammatory infiltrate and classified as normal, with nonspecific chronic or with lymphocytic gastritis. HPyl presence was diagnosed with a specialized stain (modified Giemsa) in all cases.

For DIA, images were preprocessed as recommended (30). Using open source software ilastik (31) images were segmented under three labels: inflammatory cells, epithelial cells and background (32). Segmentation probability maps were exported and evaluated using FIJI (33) (fig. 1 and 2). Each probability map was analyzed based on nuclear dimension: 5-15 microns for in-flammatory and 20-25 microns for gastric epithelial cells (34).

**Fig 1.**
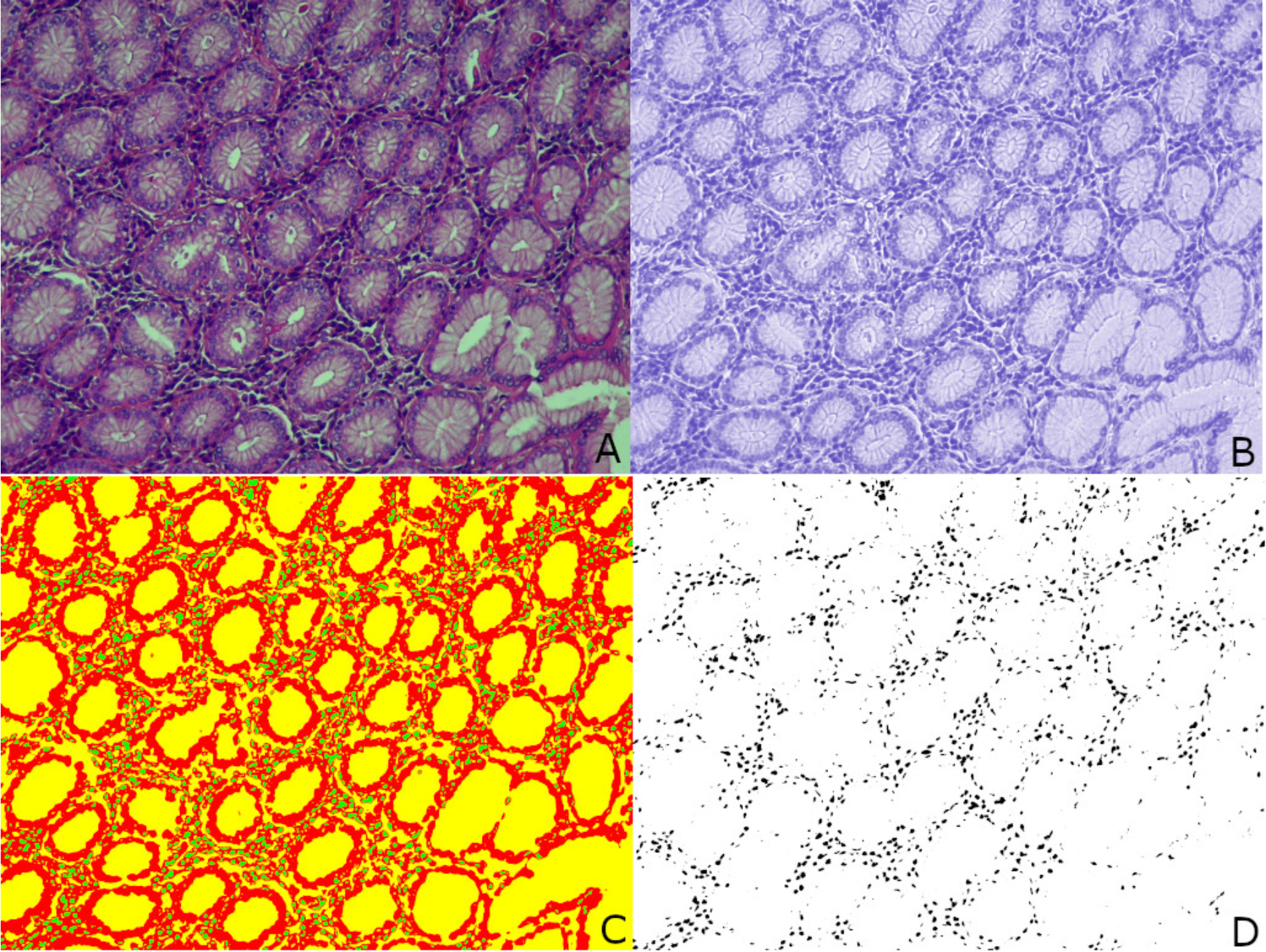
Acute Lymphocytic gastritis digital diagnostic (A. HE, training; B. channel selection, C. segmentation, in green inflammatory cells; D. binary analysis)

**Fig 2.**
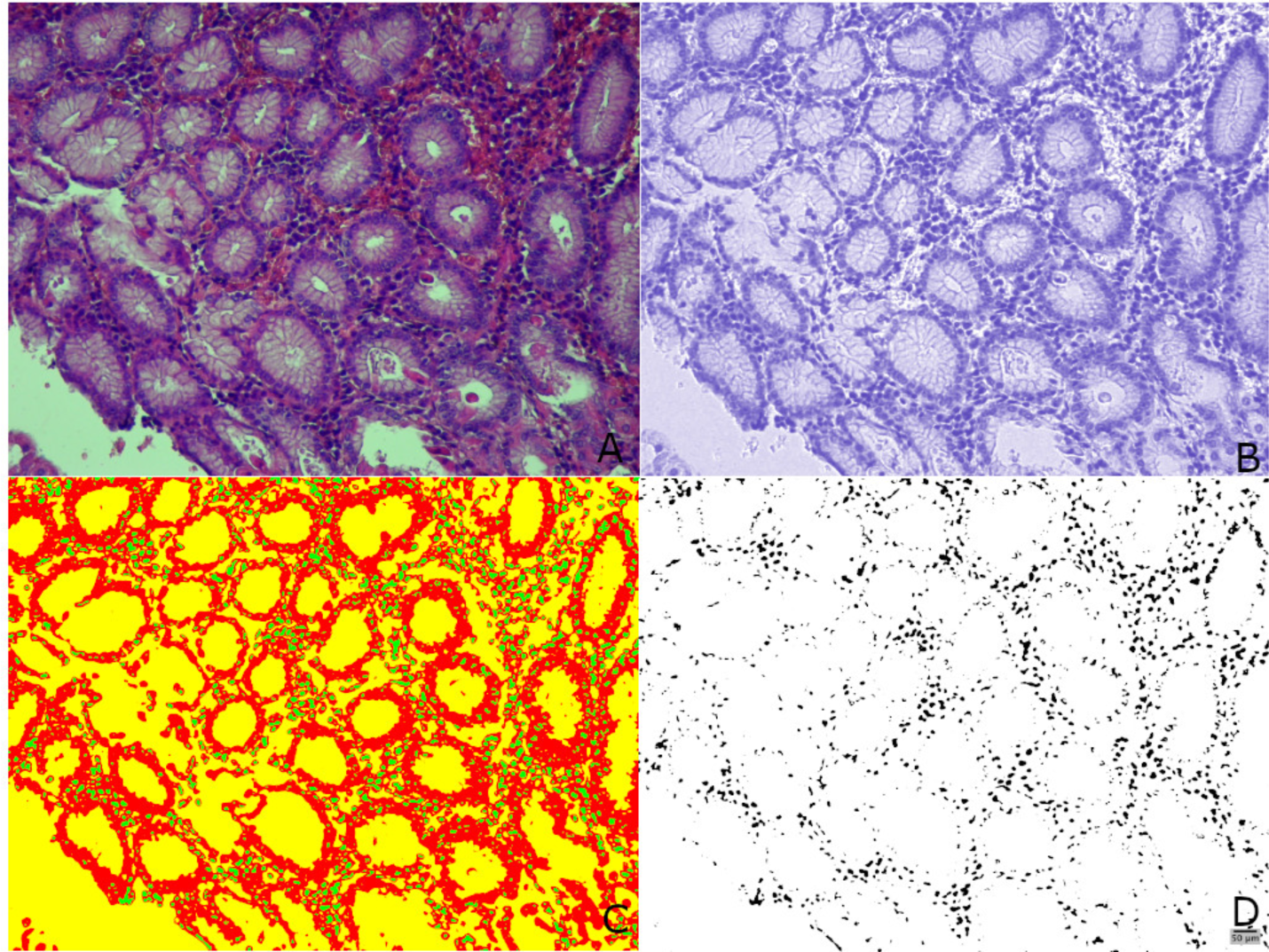
Acute lymphocytic gastritis with massive oedema, marked lymphocytosis and follicle formation.

Based on intraepithelial lymphocytes count (ratio of inflammatory over epithelial cells) each case was classified as normal, with nonspecific chronic gastritis and with lymphocytic gastritis (normal = 15/100 < lymphocytes, nonspecific chronic gastritis = under 30 lymphocytes/ 100 epithelial cells, lymphocytic gastritis = over 30/100) (35).

Discordant results were re-evaluated by the human pathologist using direct counting. The accuracy and precision of DIA classification was finally calculated (36).

The bariatric population consisted of gastric asymptomatic, severely obese patients. No adjacent pre-op HPyl diagnostic was performed. LGS was the only bariatric surgery performed and no pre-op endoscopic exam was recommended and done. Population was residing in a high-prevalence HPyl geographic area (37,38).

### Statistics

Data was stored in a standard Excel spreadsheet and was analyzed using MedCalc open source statistical software (version 19.0.7, MedCalc Software bvba, Ostend, Belgium). Basic statistic results are expressed as mean ± standard deviation. Differences between means were tested for significance with a p-value set at p<0.05.

## Ethics

Before bariatric surgery all patients provided standard written, informed consent specifically agreeing with the process and analyze of the pathology gastric samples and for participating in pathology anonymized research. The hospital IRB approved the digital analysis of images study (1122/2020).

## Results

Patients (63.21% women) had a mean age of 37.78±11.11 years and a Body Mass Index (BMI) of 41.05±3.76 kg/m2. Differences between gastric wall inflammation quantification using VAS fast scoring and DIA results are presented in table 1.

**Table 1.** Comparing gastric wall diagnostics stratification based on Visual Analogue Scale and Digital Image Analysis (*NCG=nonspecific chronic gastritis, LG=lymphocytic gastritis, IF=incidental findings). RH_HE=Hematoxylin Eosin HPyl+ detection. Giemsa=modified Giemsa HPyl detection*.

| Age Group | Nr. | Normal |  | NCG |  | LG |  | IF | Chronic Gastritis |  | HPyl+ |  |
| --- | --- | --- | --- | --- | --- | --- | --- | --- | --- | --- | --- | --- |
|  |  | VAS | DIA | VAS | DIA | VAS | DIA |  | VAS | DIA | RH_HE | Giemsa |
| <20 | 2 | 2 | 0 | 0 | 1 | 0 | 1 | 0 | 0 | 2 | 0 | 0 |
| 20-30 | 24 | 9 | 3 | 5 | 11 | 8 | 8 | 2 | 13 | 19 | 3 | 5 |
| 30-40 | 32 | 7 | 8 | 10 | 9 | 17 | 17 | 0 | 27 | 26 | 3 | 8 |
| 40-50 | 16 | 6 | 1 | 5 | 6 | 5 | 10 | 1 | 10 | 16 | 3 | 6 |
| 50-60 | 12 | 4 | 0 | 0 | 3 | 8 | 8 | 1 | 8 | 11 | 2 | 5 |
| >60 | 1 | 0 | 0 | 0 | 0 | 1 | 1 | 0 | 1 | 1 | 1 | 1 |
| Total | 87 | 28 | 12 | 20 | 30 | 39 | 45 | 4 | 59 | 75 | 12 | 25 |

Clinical-demographic data and inflammation classification for both classification methods are presented in table 2.

**Table 2.** Clinical, demographic and inflammation classification data (*RH= routine histology, DIA=digital image analysis, CG= chronic gastritis, LH= lymphocytic gastritis, BMI= body mass index, IEL=intraepithelial lymphocytes count, NA=not available, * p<0*.*0001*).

|  | Number | Sex | Age (years) | BMI (kg/m<sup>2</sup>) | IEL |
| --- | --- | --- | --- | --- | --- |
| Normal VAS | 28 | 6m / 22f | $36.5 \pm 11.3$ | $41.75 \pm 1.9^*$ | NA |
| Normal DIA | 12 | 3m / 9f | $34.8 \pm 7.03$ | $39.54 \pm 2.8$ | $14.3 \pm 0.4^*$ |
| CG VAS | 20 | 6m / 14f | $35.7 \pm 7.3$ | $38.9 \pm 1.5$ | NA |
| CG DIA | 30 | 8m / 22f | $35.86 \pm 9.6$ | $40.28 \pm 2.85$ | $25.4 \pm 0.2^*$ |
| LG VAS | 39 | 17m / 22f | $40.2 \pm 11.8$ | $37.2 \pm 2.4^*$ | NA |
| LG DIA | 45 | 19m / 26f | $40.25 \pm 11.4$ | $38.6 \pm 3.4$ | $53.1 \pm 15^*$ |

HPyl was identified using modified Giemsa in 25 cases (29%). All HPyl+ cases were graded by the human pathologist by direct counting in an area of maximal inflammation. 3 of HPyl+ cases had moderate chronic gastric inflammation, 10 had severe lymphocytosis and 12 had signs of active inflammation associated with chronic inflammation (fig.3).

**Fig 3.**
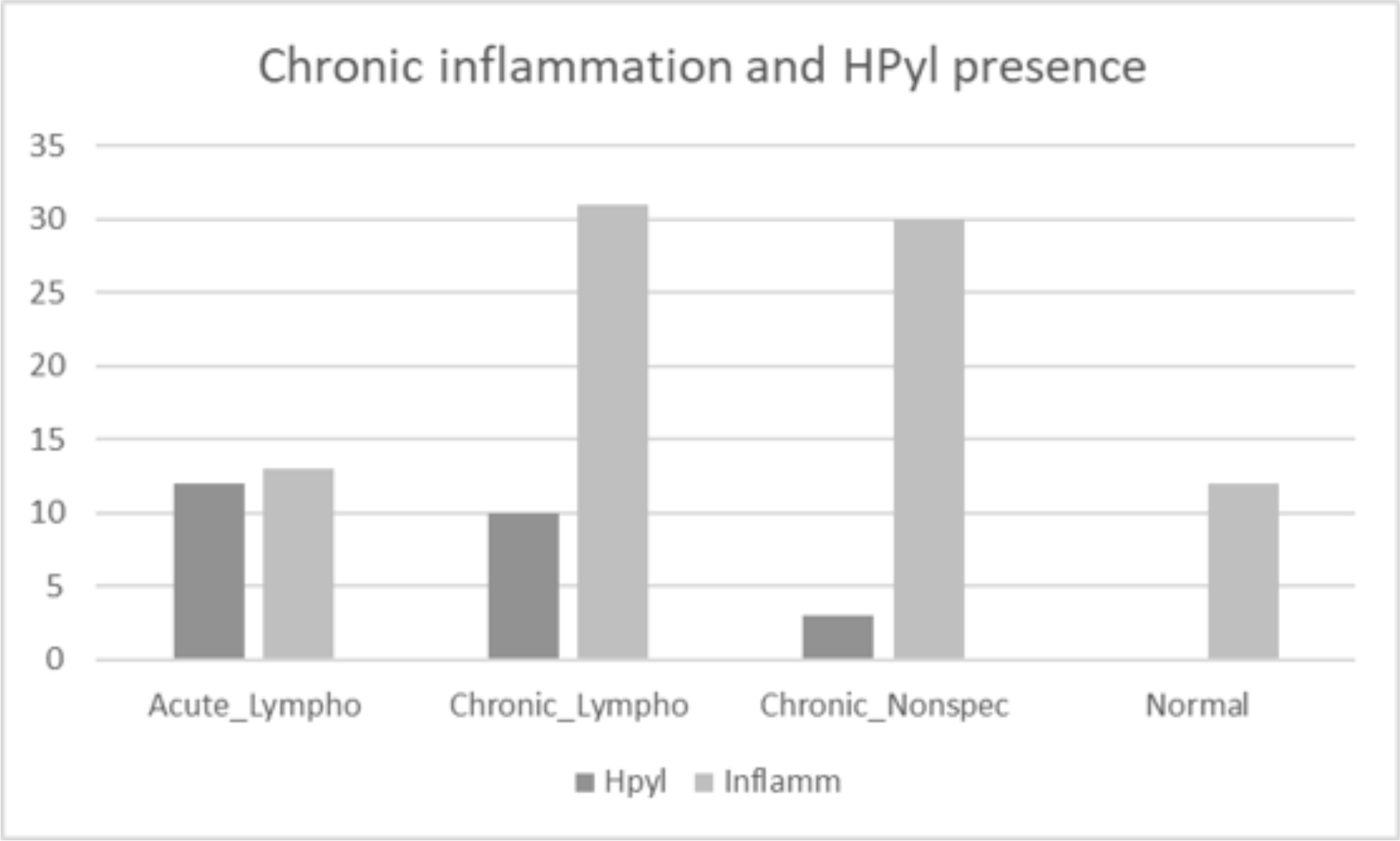
HPyl presence in relation with chronic inflammation grade.

## Discussion

Routine histology findings on normal tissue excised for therapeutic reasons are often incidental. Bariatric surgery is an exception: as many patients are infected with HPyl, the bacterium may increase the frequency and gravity of gastric wall chronic inflammation and may generate post-op complications (39). Frequently, for asymptomatic bariatric patients, gastritis diagnostic and grading are performed only on surgical samples, part of RHP exam. As the process is based on VAS fast-scoring, it is subjective, time and budget consuming. Digital quantification of the inflammatory process may help improving classification precision and produce a fast, reproducible classification.

Our study aim was to evaluate validity of manual digital quantification of chronic gastritis classification compared with human grading. Images of microscopic slides coming from 87 consecutive obese patients were prepared and analyzed using both an open source software and a routine human fast-scoring classification.

A significant difference between the IEL machine count results was observed when normal tissue, unspecified gastritis and lymphocytic gastritis groups were compared. Previously published data concerning IEL value in gastritis classification performed using direct microscopic cell count (40,41) reported similar differences.

In our cohort, HPyl presence was diagnosed in 22 cases of lymphocytic gastritis (48%) and in only 3 cases of nonspecific chronic gastritis (10%). Within the lymphocytic group, 12 cases had signs of acute inflammatory activity (IEL mean of 60.3±18). 10 HPyl+ cases did not show signs of acute inflammatory reaction (IEL mean of 49.4±13). For the 3 cases detected in the chronic nonspecific gastritis group the mean IEL was 24 (±0.2).

DIA quantification results were concordant with human VAS in 47 cases (54%). All 40 discordant cases were re-examined by the surgical pathologist by direct counting in an area of maximal inflammation. DIA results were true positive in 27 cases, true negative in 4 cases; in 3 cases results were false positive and in 6 cases were false negative. Slides technical artifacts (that did not generate any difficulty for human scoring) were responsible for all 6 false negative results.

DIA demonstrated a robust accuracy (89%) and precision (96%) when compared with visual analogue fast scoring.

In our general surgical pathology department, more than 60% of time and budget is allocated to routine histology diagnostic. Of it, 5-7.5% of all RHP exams are performed on normal gastric tissue sampled after laparoscopic gastric sleeve from gastric asymptomatic, severely obese patients and is focusing on gastritis and HPyl diagnostics. Gastritis classification is performed using VAS and may be subjective. Digital quantification of inflammation, as an alternative to human scoring, was fast, reproducible and worked well in classification of moderate and severe lymphocytic infiltrates cases. Mild lymphocytic infiltrates classification generated most of differences between human fast scoring and machine analysis.

A well-known limitation of DIA diagnostic in benign histopathology (seen in our study as well) is given by fixing, cutting and staining techniques. Better slide preparation will improve DIA diagnostic validity by eliminating false negative cases. Another limitation of the method was the selection of LGS cases (fundus gastric tissue only). Both chronic inflammation and HPyl presence may differ at the fundic level from other gastric areas. The use of bariatric patients without any gastric symptomatology allowed a homogenous patient population and LGS is the only laparoscopic procedure performed in our centre on asymptomatic patients. The manual segmentation process (an important aspect of digital analysis based on supervised training) can be as subjective as the human fast-scoring process.

To our knowledge, this is a first study that evaluates the utility of digital image analysis in routine histopathology. Based on our data we consider that digital image analysis can help routine diagnostic in terms of time and reproducibility when normal tissue is examined even when a manual analysis is performed. Automation of the whole process may increase the performance of the diagnostic but will also increase operating costs.

## Data Availability

raw data is publicly available at https://data.mendeley.com/datasets/j3hpsvbgrb/draft?a=555a000b-7973-4a23-a47e-3dd2f58401ce

## Author Statement

CV and BG conceived the idea, CV performed the histologic evaluation, MG performed the digital image acquisition, digital analysis and first draft conception, all authors reviewed the final article.

## References

1. M Stolte, A Meining. The updated Sydney system: Classification and grading of gastritis as the basis of diagnosis and treatment. Can J Gastroenterol 2001;15(9):591–59

2. Saafan T., Basah M., Ansari W, Karam M., Histopathological changes in laparoscopic sleeve gastrectomy specimens: prevalence, risk factors, and value of routine histo-pathologic examination Obesity Surgery 2017 27(7): 1741–1749 http://dx.doi.org/10.1007/s11695-016-2525-1

3. Crowe SE. Helicobacter infection, chronic inflammation, and the development of malignancy. Curr Opin Gastroenterol. 2005;21:32

4. Meining A, Stolte M, Hatz R, et al. Differing grade and distribution of gastritis in Helicobacter pylori associated diseases. Virchows Arch 1997;431:11–5

5. Toulaymat M, Marconi BS, Garb MS, et al, Endoscopic biopsy pathology of Helicobacter pylori gastritis comparison of bacterial detection by immunohistochemistry and genta stain. Arch Pathol Lab Med. 1999;123:778–81

6. Sipponen P., Price A., The Sydney System for classification of Gastritis 20 years ago, Journal of Gastroenterology and Hepatology 26 (2011) Suppl. 1; 31–34, doi:10.1111/j.1440-1746.2010.06536.x

7. Henriques U., Histological Technique in Routine Histopathology: an opinion, Pathology Research and Practice, 171 3–4, 1981, 417-422

8. Netser J., Robinson R., et al, Valued based pathology, a cost-benefit analysis of the examination of routine and non-routine tonsil and adenoid specimens, Am J Clin Pathol 1997;108:158–165

9. Cook IS, Fuller CE. Does histopathological examination of breast reduction specimens affect patient management and clinical follow-up? J Clin Pathol 2004;57:286–289

10. Lemarchand N, Tanne F, Aubert M., et al, Is routine pathologic evaluation of hemorrhoidectomy specimens necessary? Gastroenterol Clin Biol 2004 28:659–661

11. Pitre, E. Routine Ordering of Primary Pathology Examinations in Canada. Ottawa: Canadian Agency for Drugs and Technologies in Health; 2014. (Environmental Scan; Issue 46

12. Matthyssens LE, Ziol M, Barrat C, Champault GG (2006) Routine surgical pathology in general surgery. Br J Surg 93:362–368

13. Ramraje SN, Pawar VI, Routine Histopathologic Examination of Two Common Surgical Specimens Appendix and Gallbladder: Is It a Waste of Expertise and Hospital Resources? Indian J Surg 2014, 76(2):127–130, DOI 10.1007/s12262-012-0645-y

14. Nguyen NT, Root J, Zainabadi K, et al, Accelerated growth of bariatric surgery with the introduction of minimal invasive surgery. Arch Surg 2005;140(12) 1198-202; doi: 10.1001/archsurg.140.12.1198

15. Buchwald H, Varco R. Metabolic surgery. New York: Grune & Stratton; 1978

16. Hansen SK, Pottorf BJ, Hollis Jr HW, et al, Is it necessary to perform full pathologic review of all gastric remnants following sleeve gastrectomy?, The American Journal of Surgery (2017), doi: 10.1016/j.amjsurg.2017.06.029

17. Vrabie CD, Gangal MD, Clinical effectiveness of routine pathological exam after bariatric laparoscopic sleeve gastrectomy J Clin Exp Pathol 2018, 8:5 DOI: 10.4172/2161-0681.1000357

18. Brownlee A., Bromberg E., Roslin M., Outcomes in Patients with Helicobacter pylori Undergoing Laparoscopic Sleeve Gastrectomy. OBES SURG (2015) 25:2276–2279 DOI 10.1007/s11695-015-1687-6

19. Sipponen P. Helicobacter pylori: a cohort phenomenon. Am J Surg Pathol. 1995;19 (Suppl 1):S30–S36

20. Dixon MF, Genta RM, Yardley JH. Classification and grading of gastritis. The Updated Sydney System. International Workshop on the histopathology of gastritis, Houston 1994. Am J Surg Pathol 1996;20:1161–81

21. Prewitt JMS, Mendelsohn ML., The Analysis of Cell Images, Annals of the New York Academy of Sciences, 1965, 128: 1035–1053

22. Gurcan M., Boucheron L., et al Histopatological Image Analysis, a review, IEEE Rev Biomed Eng. 2009 ; 2: 147–171. doi:10.1109/RBME.2009.2034865

23. Bhattacharjee S., Mukherjee J., et al, Review on Histopathological Slide Analysis using Digital Microscopy, International Journal of Advanced Science and Technology Vol.62, (2014), pp. 65–96 http://dx.doi.org/10.14257/ijast.2014.62.06

24. Kourou K, Exarchos T.P., et al, Machine learning applications in cancer prognosis and prediction, Computational and Structural Biotechnology Journal, vol. 13, pp. 8–17, 2015.

25. Snead D R J, Tsang Y-W, Meskiri A, et al, Validation of digital pathology imaging for primary histopathological diagnosis Histopathology 2016 68, 1063–1072. DOI: 10.1111/his.12879

26. Fuchs TJ, Buhmann JM. Computational pathology: challenges and promises for tissue analysis. Comput. Med. Imaging Graph. 2011; 35(7-8):515–530

27. Djuric U., Zadeh G., et al, Precision histology: how deep learning is poised to revitalize histomorphology for personalized cancer care, npj Precision Oncology (2017) 1:22 ; doi:10.1038/s41698-017-0022-1

28. Zarella MD, Bowman D, et al A Practical Guide to Whole Slide Imaging: A White Paper From the Digital Pathology Association. Archives of Pathology & Laboratory Medicine: 2019, 143, 2, 222–234 https://doi.org/10.5858/arpa.2018-0343-RA.

29. Vrabie CD, Cojocaru M, Waller M, Sindelaru R, Copaescu C, The main histopathological gastric lesions in obese patients who underwent sleeve gastrectomy. Dicle Med J. 2010;37(2):97–103

30. Haubold C., Schiegg M., Kreshuk A., et al, Segmenting and Tracking Multiple Dividing Targets Using ilastik. In: De Vos W., Munck S., Timmermans JP. (eds) Focus on BioImage In-formatics. Advances in Anatomy, Embryology and Cell Biology, 2016 vol 219. Springer

31. Berg, S., Kutra, D., Kroeger, T. et al. ilastik: interactive machine learning for (bio)image analysis. Nat Methods 16, 1226–1232 (2019) doi:10.1038/s41592-019-0582-9

32. Sommer C., Straehle C., ilastik: Interactive Learning and Segmentation kit, Heidelberg Collaboratory for Image Processing (HCI), University of Heidelberg January 24, 2011

33. Schindelin, J.; Arganda-Carreras, I. & Frise, E. et al. (2012), “Fiji: an open-source platform for biological-image analysis”, Nature methods 9(7): 676-682, PMID 22743772, doi:10.1038/nmeth.2019

34. Bergman R., Afifi A., Anatomy digital library https:cAnatomy/Section04/Section04.shtml

35. Haot J, Hamichi L, et al Lymphocytic gastritis: a newly described entity: a retrospective endoscopic and histological study. Gut 29: 1258-1264, 1988

36. Eusebi P., Diagnostic Accuracy Measures, Cerebrovasc Dis 2013;36:267–272 DOI: 10.1159/000353863

37. Olar L., Mitrut P., Evaluation of Helicobacter pylori infection in patients with eso-gastro-duodenal pathology, RomJMorpholEmbryol 2017, 58(3) 809–815,

38. Danciu M, Simion L, et al, The role of histological evaluation of Helicobacter pylori infection in obese patients referred to laparoscopic sleeve gastrectomy Rom J Morphol Embryol 2016, 57(4):1303–1311

39. Mocanu V., Dang JT et al, The Effect of Helicobacter pylori on Postoperative Outcomes in Patients Undergoing Bariatric Surgery: a Systematic Review and Meta-analysis, Obesity Surgery https://doi.org/10.1007/s11695-017-3024-8

40. Haot J., Jouret A., Lymphocitic gastritis - prospective study of its relationship with variolo-form gastritis, Gut 1990, 31, 282–285

41. Dixon MF, Wyatt JI, Burke DA, et al. Lymphocytic gastritis: relationship to Campylobacter pylori infection. J Pathol., 1988, 154:125–132.

